# Community Perception and Attitude towards people with schizophrenia among residents of Arba Minch Zuria District, Arba Minch Health and Demographic Surveillance System sites (AM-HDSS), Ethiopia. Cross-section study

**DOI:** 10.1101/2019.12.18.19015271

**Authors:** Negussie Boti, Sultan Hussen, Gistane Ayele, Abera Marsha, Selamawit Gebeyehu, Mekidm Kassa, Tesfaye Feleke, Gebremaryam Temesgen

## Abstract

**Background:** Mental disorders are one of the public health issues throughout the worldwide. Among people with mental disorders, more than 21 million people have schizophrenia. However, there is scarcity of information about perception and attitude of the community toward people with schizophrenia in Ethiopia including the study area.

**Objective:** To assess the community perception and attitude towards people with schizophrenia among residents of Arba Minch Zuria District, Arba Minch Health and Demographic Surveillance System sites (AM-HDSS), Southern Ethiopia

**Methods:** A community-based cross-sectional study was conducted among 617 randomly selected residents of Arba Minch Zuria District, AM-HDSS Site. The data was collected using structured, pre-tested and interviewer-administered questionnaires. Descriptive statistics including frequencies, means, and median were performed. Binary and multivariable logistic regression analyses were performed to identify factors affecting community attitude.

**Results:** The finding of this study showed that, among study participants 469(76%) have a good perception towards people with schizophrenia and 390(63.2%) of respondents have positive attitudes towards people living with schizophrenia. Talkativeness and self-neglect behaviors were the most commonly mentioned manifestation of schizophrenia. Substance misuse and head injury were the most perceived cause of schizophrenia. Spiritual/traditional methods were preferred place for the treatment of schizophrenia. Age of respondents, sex of respondent; educational status and perception toward people with schizophrenia were significantly associated with positive attitude towards people with schizophrenia.

**Conclusions:** Among our rural adults nearly half of the study participants have a negative attitude towards people with schizophrenia. Therefore, giving special attention to females, youths, uneducated and individuals who have poor perception toward people living with schizophrenia is crucial. Also, future mental health promotion activities should focus on cause and common manifestation of schizophrenia to improve the community attitude toward people with schizophrenia

## Introduction

Mental health disorders are health conditions that show abnormal behavior and thinking associated with distress and problems functioning in family activities [1,2]. Schizophrenia is the most common mental illness that affects educational and occupational performance [3,4].

Globally, Schizophrenia affects more than 21 million people that affect males more than 12 million, than females more than 9 million[3]. Africa accounts for 0.5 % of the world population[4]. In Ethiopia, schizophrenia was ranked in the top ten causes of morbidity from the total burden of disease [5].

Even though schizophrenia was one of the public health problems in Africa the majority of those patients did not get treatment due to neglected in Africa’s health especially in Ethiopia[6]. The “treatment gap” – the proportion of people with mental illness who don’t get treatment – ranges from 75% in South Africa to more than 90% in Nigeria[6]. In Ethiopia, among people living with schizophrenia only 10% ever receive effective care [4].

Evidence shows that one of the barriers to seeking help for people with schizophrenia was negative attitude and perception concerning mental health are influenced by a lack of knowledge as well as a mix of traditional and modern views [7,8]. In Africa, only 35% of people living with schizophrenia were taken from modern psychiatric treatment. One of the reasons for this was due to an unacceptable belief on the causes of mental illnesses and the type of treatment sector[9]. In Sub-Saharan Africa stigmatizing attitudes towards the mentally ill exists, especially in the rural areas with scant research in other countries and mirrored by the experienced stigma reported by the patients [10,11].

In Ethiopia, mental health has been neglect that are not given attention for a long time, thus most people use traditional methods for treating mental illness[5,12]. Study conducted in different part of Ethiopia reveals that due to lack of available local means psychotic treatment many families keep the people living with schizophrenia at home under restraint until they are no longer aggressive and violent once the disorderly behavior is over they will be released from restraint and many of them become wanderers and homeless. As a result, mental health services are not given due priority and the needs of people for mental health care are not met [5,12,13].

Most Ethiopians’ perceived that schizophrenia is an illness caused by supernatural forces. This influences the attitude towards mental illness and help-seeking associated with mental health problems [13]. Also, several factors are associated with the community perception and attitude related to schizophrenia which is mainly socio-demographic including gender, age, religion, income and education[13-15]. However, little is known about the perception and attitude of the community regarding people living with schizophrenia in Ethiopia including the study area [13-15]. Therefore, an in-depth understanding of community perception and attitude towards people with living with schizophrenia may help to develop interventions that may contribute to an enabling environment to fully access and utilize mental health services. So, this study aimed to assess the recent status of community perception and attitude towards people with schizophrenia among residents of Arba Minch Zuria District, AM-HDSS Site, and Southern Ethiopia.

## Methods

### Study Area and Period

This study was conducted in the nine kebeles of Arba Minch Zuria districts included under Arba Minch Health and Demographic Surveillance System sites (AM-HDSS). Arba Minch town was capital city and located at 450km from Addis Ababa and 275km from regional town, Hawassa. The district was bordered on the south by the Dirashe special woreda, on the west by Bonke, on the north by Dita and Chencha, on the northeast by Mirab Abaya, on the east by the Oromia Region, and on the southeast by the Amaro special woreda. Arba Minch was found at an altitude of 130 above sea level with the average temperature of 29°C. Based on the 2007 Census conducted by the CSA, this district has a total population of 164,529, of whom 82,199 are men and 82,330 women. Arba Minch Zuria district has a total of 31 kebeles with three different climatic zones, high land, midland and lowland, among which 9 kebeles are used as HDSS. The study was conducted from January 10 to 24, 2019**(Figure 1)**

**Figure 1:**
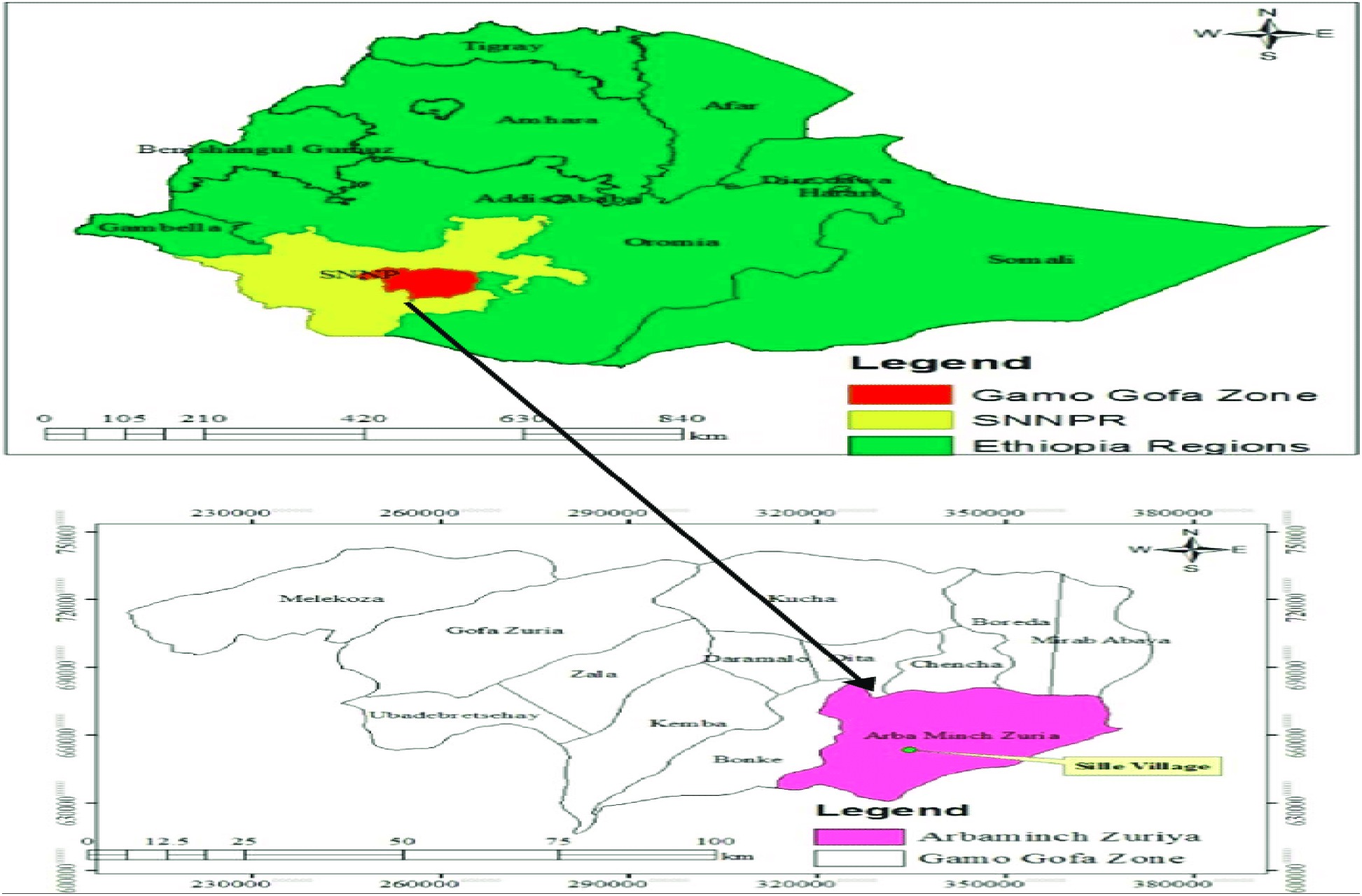
Map of Study area, Arba Minch Zuria District, AM-HDSS Site, Ethiopia, 2019

### Population

All households in the AM-HDSS Site was the source population and all head of households (or their spouse) who were living in AM-HDSS during the data collection period and full fill inclusion criteria were the study population

### Eligibility criteria

All residents who were 18 years old and above living for at least 6 months in the study area during the time of the study were included in this study. Those who were critically ill during the data collection period were excluded.

### Sample size and sampling technique

The sample size for this study was determine by using single population proportion formula considering the following assumptions: the proportion of the community who had poor perception towards mental illness was 37.3 %[14], 95% confidence level (1.96), 4%, the margin of error, and 10% nonresponse rate. Therefore, the minimal sample size required for this study was **617**. To select the study participants, we used a sampling frame of the AM-HDSS database. Then, based on the total number of households living in each kebele, study participants were proportionally allocated. Then using a simple random sampling method the number of study participants was selected using computer-generated random numbers.

### Measurement

#### Attitude toward people with schizophrenia

It refers to a community feeling of expression towards mental illness. It was measured based on 13 attitude related questions. To categorized community attitude towards people with schizophrenia, we used the Demarcation threshold formula. Those who score ≥ 39 were considered to have a good attitude towards people with schizophrenia [5,13]. Demarcation threshold formula

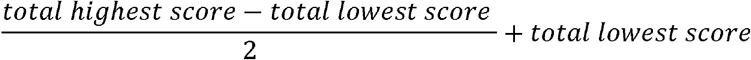

#### Community Perception toward people with schizophrenia

It refers to the level of identification, and interpretation of sensory information to represent and understand the presented information. It was assessed by 9 items of five Likert scale perception related questionnaires. To categorized community perception towards people with schizophrenia, we used the Demarcation threshold formula. Those who score ≤27 for 9 items of five semantic differential scales were considered having a good perception towards people with schizophrenia [13,14].

### Data collection procedure

The data was collected by using pre-tested structured interviewer-administered questionnaires which was developed by reviewing different literatures [13-15]. The questionnaires were asked after reading the vignette case. As recommended by WHO a case vignette we used modified vignettes to assess community perception toward people living with schizophrenia using 9 items of five Likert scale perception related questionnaires from 1 to 5 (1 = strongly agree, 2 = agree, 3 = neutral, 4 = disagree, and 5 = strongly disagree). To assess community attitude towards people living with schizophrenia we used 13 items of five-point Likert scale from 1 to 5 (1 = strongly agree, 2 = agree, 3 = neutral, 4 = disagree, and 5 = strongly disagree). This tool also has been previously pretested and validated in Ethiopia and other Sub-Saharan Africa [13,14,16].

### Case of Vignettes for schizophrenia

**“**During the last six months, one of your friends has changed. He withdraws from his coworkers and friends more and more. He keeps out of everybody’s way. Contrary to his former habits, he does not take care of his appearance any longer and seems to neglect himself increasingly. He seems to be anxious and agitated. He reports to be convinced that people are able to read other people’s thoughts, and that they are also able to influence these thoughts; but he would not yet know who is controlling his thoughts. He even hears these people talking to him and giving him orders. Sometimes, they speak to one another and mock him. In his apartment, the situation is particularly bad. There he feels threatened and terribly scared. He has not been at home for a week and hid in a hotel, which he has not dared to leave.” The tool was first developed in the English language and translated to Amharic language then back to the English language to check the consistency.

### Data quality control

To maintain data quality was ensured during instrument development, collection, coding, and analysis. Training was given to the supervisors and data collectors. The data collection tool was pre-tested on 5% of the study participants in Mirab Abaya, before the actual data collection. During data collection, the collected questionnaire was checked for its completeness every day by investigators and supervisors

### Data processing and analysis

Each questionnaire was checked for completeness. Data was collected using ODK software. The collected data was cleaned and edited and analysis by using STATA version 14.0. A descriptive analysis was conducted. Then, a binary logistic regression analysis was performed for each independent variable and outcome of interests to identify independent predictors. Upon the completion of the bivariate logistic regression analysis, variables with p-value <0.25 were selected for the multivariable analysis. Context and previous studies were also being considered to make a variable candidate for multivariable analysis. To assess the strength of association the p-value ≤ 0.05 was used. A multi-collinearity assumption and goodness of fit test was measured using the variance inflation factor (VIF), Hosmer and Lemeshow test respectively.

## Results

### Socio-demographic characteristics of the study participants

A total of 617 individuals participated in this study with a response rate of 100%. Among the total 617 respondents, 324(52.5%) were male and 559(90.6%) of them ever been married. The mean (±SD) age of respondents was 35.61 (±10.52) years. The majority of respondents were 507(82.2%) were Gamo by ethnicity and 423(68.6%) were protestant Christian in their religion. Regarding their educational status, 312(50.6%) of the participants were completed primary school and 615(99.7%) were rural residents (Table 1).

**Table 1:** Sociodemographic characteristics of respondents residing in AMU-HDSS, Southern, Ethiopia, 2019. (N = 617)

| Variables | Category | Frequency | Percent |
| --- | --- | --- | --- |
| Age group | 18-24 | 170 | 27.60 |
|  | 25-44 | 252 | 40.80 |
| | $\geq 45$ | 195 | 31.60 |
| Age (mean ± SD) | 35.61 ± 10.52 | | |
| Sex | Male | 324 | 52.50 |
|  | Female | 293 | 47.50 |
| Religion | Orthodox | 188 | 30.5 |
|  | Protestant | 423 | 68.6 |
|  | Muslim | 2 | 0.3 |
|  | Other* | 4 | 0.6 |
| Ethnicity | Gamo | 507 | 82.2 |
|  | Gofa | 3 | 0.5 |
|  | Zayse | 52 | 8.4 |
|  | Woliata | 51 | 8.3 |
|  | Oromo | 3 | 0.5 |
|  | Other** | 1 | 0.2 |
| Educational status | No formal education | 224 | 36.3 |
|  | Primary(1-8) | 312 | 50.6 |
|  | Secondary(9-12) | 64 | 10.4 |
|  | College and above | 17 | 2.8 |
| Residence | Urban | 2 | 0.3 |
|  | Rural | 615 | 99.7 |
| Marital status | Married | 559 | 90.6 |
|  | Single | 30 | 4.86 |
|  | Divorced | 28 | 4.54 |
| Occupational status | Farmers | 311 | 50.4 |
|  | Merchant | 52 | 8.4 |
|  | Governmental Employer | 16 | 2.6 |
|  | Daily laborer | 98 | 15.9 |
|  | Students | 140 | 22.7 |
| Family history of mental illness | Yes | 45 | 7.3 |
|  | No | 572 | 92.7 |
| Wealth index | Low | 202 | 32.70 |
|  | Moderate | 210 | 34.02 |
|  | High | 205 | 33.20 |

\*traditional \*\*Kore

### Sources of information and exposure to people with schizophrenia-related problems

Around 558(90.4%) of the participants ever heard about schizophrenia. The main sources of information for schizophrenia were mass media which accounts, 399(48%) followed by religious institutions 113(13.6%) of the respondents. Regard to perception about schizophrenia, among respondents 401(71.86%) list the possible cause of schizophrenia, among those majority 367(91.52%) and 275(68.58%) of perceived that the cause of schizophrenia is substance misuse and head injury cause illness respectively. Also, 341(61.11%) of respondents mentioned that preferential treatment for people with schizophrenia was spiritual/traditional **(Table 2)**.

**Table 2:**
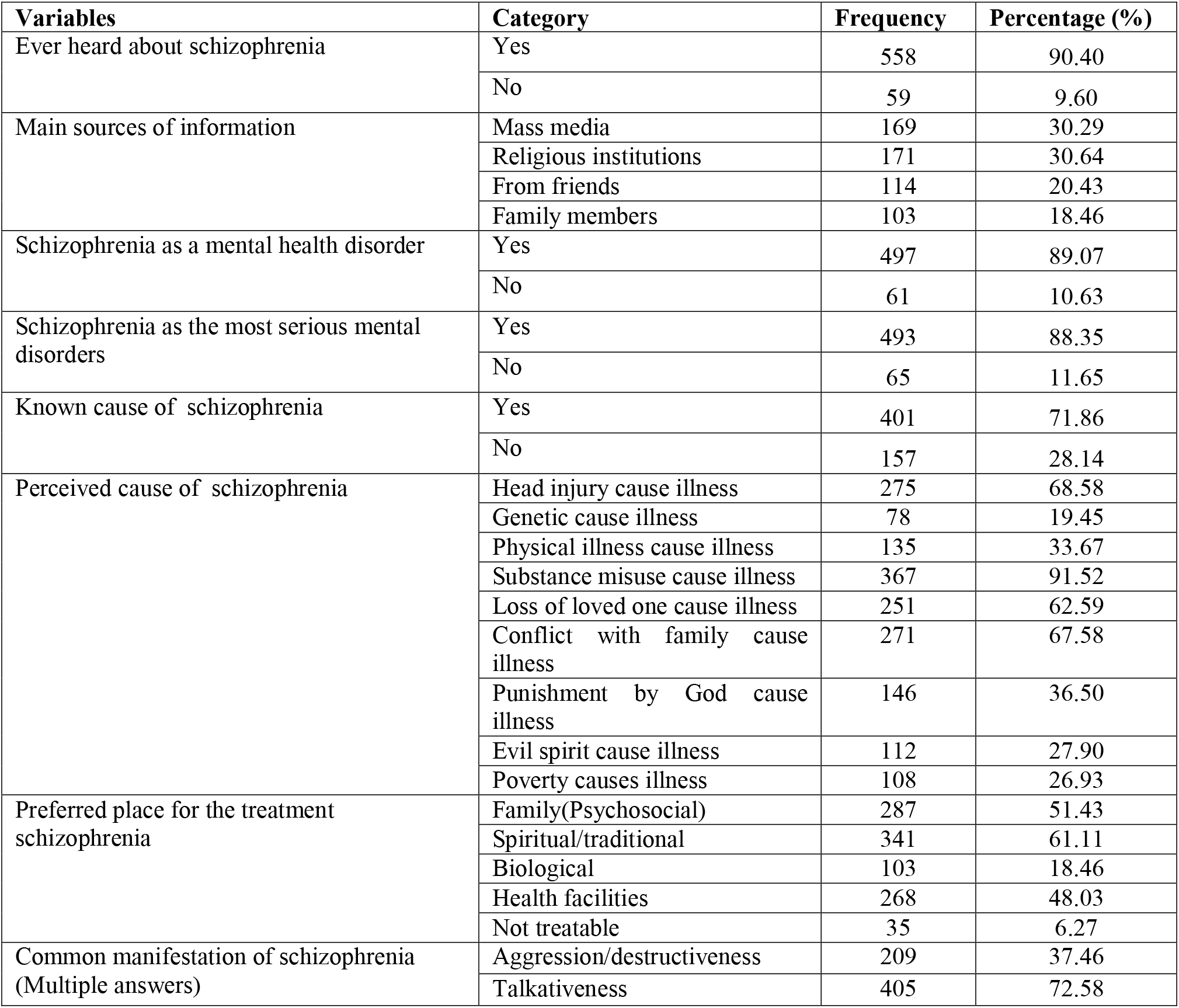
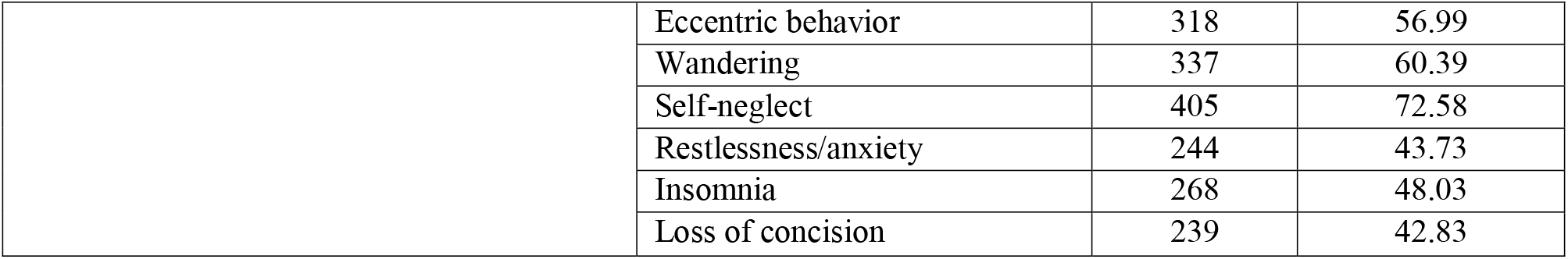
Source of information about schizophrenia among respondents residing in AMU-HDSS, Southern, Ethiopia, 2019. (N = 617)

### Community perception and attitude towards people with schizophrenia

The finding of this study found that 76.01 % (95% CI: 72.6 – 79.4%) of respondents have a good perception about people with schizophrenia and 63.21% (95% CI: 59.4– 67%) of respondents have a positive attitude towards people with schizophrenia **(Table 3)**.

**Table 3:** Community perception and attitude towards people with schizophrenia among residents of Arba Minch Zuria District, AM-HDSS Site, Southern Ethiopia

| **Variables** | **Category** | **Frequency** | **Percentage** |
| --- | --- | --- | --- |
| Community perception towards people with schizophrenia | Good Perception | 469 | 76.01 |
|  | Poor Perception | 148 | 23.99 |
| Community attitude towards people with schizophrenia | Positive attitude | 390 | 63.21 |
|  | Negative attitude | 227 | 36.79 |

\*Good perception means who score $\leq 27$ and Positive attitude means who score $\leq 39$

### Community attitude towards people with schizophrenia

The finding of this study found that 243(39.4 %) of respondents agree that as a person shows signs of schizophrenia, he or she should be hospitalized, more than half (55.9%) agreed that schizophrenia is caused by substance abuse. Nearly one-fourth (22.4%) agreed that if treated and medicated, people with depression can function fairly typically in society. Also half (49.1%) of respondents agree individuals with schizophrenia are simply weak-willed, unmotivated people. More than half (70.5%) of respondents agree that people with schizophrenia are dangerous and half (49.4%) of respondents disagree individuals with schizophrenia do not need medication; they just need to change their thought processes and behaviors. Also nearly half (49.6 %) of respondents disagree that people with depression should have the same educational, occupational, and social opportunities as ‘‘normal’’ individuals. More than half (65.8%) of respondents agree people with schizophrenia have behavioral patterns that are abnormal and nearly one-sixth (12.6%) of respondents agree most people fear people with schizophrenia (Table 4).

**Table 4:** Responses to the item of community attitude towards people with schizophrenia subscales among residents of Arba Minch Zuria District, AM-HDSS Site, Ethiopia, 2019(N=617)

| Items | Strongly Agree | Agree | Neutral | Disagree | Strongly disagree |
| --- | --- | --- | --- | --- | --- |
|  | N (%) | N (%) | N (%) | N (%) | N (%) |
| Do you agree as soon as a person shows signs of schizophrenia, he or she should be hospitalized? | 88(14.3) | 243(39.4) | 95(15.4) | 171(27.7) | 20(3.2) |
| Do you agree schizophrenia is caused by substance abuse? | 167(27.1) | 345(55.9) | 13(2.1) | 89(14.4) | 3(0.5) |
| Do you agree individuals with schizophrenia are simply weak-willed, unmotivated people? | 91(14.7) | 303(49.1) | 88(14.3) | 131(21.2) | 4(0.6) |
| Do you agree if treated and medicated, people with schizophrenia can function fairly typically in society? | 127(20.6) | 246(39.9) | 101(16.4) | 138(22.4) | 5(0.8) |
| Do you agree people with schizophrenia are dangerous? | 115(18.6) | 435(70.5) | 26(4.2) | 41(6.6) | 0.00 |
| Do you agree with a person with schizophrenia's social problems is their fault because they isolate themselves from others? | 76(12.3) | 294(47.6) | 106(17.2) | 136(22) | 5(0.8) |
| Do you agree genetics are the primary factor in the development of schizophrenia? | 49(7.9) | 70(11.3) | 81(13.1) | 370(60) | 47(7.6) |
| Do you agree most people with schizophrenia are poor? | 24(3.9) | 45(7.3) | 46(7.5) | 334(54.1) | 168(27.2) |
| Do you agree individuals with schizophrenia do not need medication; they just need to change their thought processes and behaviors? | 29(4.7) | 75(12.2) | 108(17.5) | 605(49.4) | 100(16.2) |
| Do you agree individuals with schizophrenia are victims of their disease and should be treated with sympathy? | 81(13.1) | 288(46.7) | 79(12.8) | 163(26.4) | 6(1.0) |
| Do you agree people with schizophrenia have abnormal behavioral patterns? | 136(22) | 406(65.8) | 35(5.7) | 38(6.2) | 2(0.3) |
| Do you agree people with schizophrenia should have the same educational, occupational, and social opportunities as “normal” individuals? | 57(9.2) | 82(13.3) | 120(19.4) | 306(49.6) | 52(8.4) |
| Do you agree most people fear people with schizophrenia? | 53(8.6) | 78(12.6) | 119(19.3) | 314(50.9) | 53(8.6) |

### Factors associated with community attitude towards people with schizophrenia

After controlling confounding effect variables like age, sex, educational status, and perception towards people living with schizophrenia were significantly associated with a positive attitude towards people living with schizophrenia in multivariable logistic regression. However, marital status, wealth index, family history of mental illness and having information about schizophrenia were not significantly associated with community attitude towards people with schizophrenia in multivariable logistic regression.

The finding of this study reveals that respondents whose age between 18–24 years were 2.03 times more likely to have a positive attitude towards people with schizophrenia compared to older age ***[AOR=2.03, 95% CI: 1.21,3.40]***.In addition, those females were more likely to have a positive attitude compared to males ***[AOR=2.32, 95% CI: 1.58,3.41]***. Moreover, having poor perception towards people with schizophrenia was 4.95 times more likely to have a positive attitude towards people with schizophrenia compared to those who were good perception towards people with schizophrenia ***[AOR=4.95, 95% CI: 3.25 – 7.54]*. (Table 5)**

**Table 5:** Factors associated with community attitude towards people with schizophrenia among residents of Arba Minch Zuria District, AM-HDSS Site, Southern Ethiopia, 2019

| Variables | Categories | Attitude towards people with schizophrenia |  | COR [95% CI] | AOR [95% CI] | P-value |
| --- | --- | --- | --- | --- | --- | --- |
|  |  | Positive N (%) | Negative N (%) |  |  |  |
| Age in years | 18–24 | 93(54.7) | 77(45.3) | 1.96(1.27,3.01) | **2.03(1.21,3.40)** | **0.007\*** |
|  | 25–44 | 160(63.5) | 92(36.5) | 1.36(0.91,2.03) | 1.23(0.78,1.93) | 0.370 |
|  | ≥45 | 132(70.3) | 58(29.5) | 1.00 | 1.00 |  |
| Sex | Male | 233(71.9) | 91(40.1) | 1.00 | 1.00 |  |
|  | Female | 157(53.6) | 136(46.4) | 2.22(1.59,3.09) | **2.32(1.58,3.41)** | **0.001\*** |
| Level of Education | No formal education | 142(63.4) | 82(36.6) | 3.02(1.57,5.80) | 2.75(1.33,5.70) | **0.007\*** |
|  | Primary education | 180(57.7) | 132(42.3) | 3.84(2.03,7.23) | 3.72(1.87,7.39) | **0.001\*** |
|  | Secondary and above | 68(84.0) | 13(16.0) | 1.00 | 1.00 |  |
| Marital status | Married | 349(62.4) | 210(37.6) | 1.00 | 1.00 |  |
|  | Single | 41(70.7) | 17(29.3) | 0.69(0.38,1.24) | 0.73(0.38,1.42) | 0.356 |
| Family History of MI | Yes | 29(64.4) | 16(35.6) | 1.00 | 1.00 |  |
|  | No | 361(63.1) | 211(36.9) | 1.06(0.56,1.99) | 1.30(0.65,2.62) | 0.462 |
| Ever heard about MI | Yes | 352(63.1) | 206(36.9) | 1.06(0.61,1.85) | 1.05(0.56,1.98) | 0.871 |
|  | No | 38(64.4) | 21(35.6) | 1.00 | 1.00 |  |
| **Community perception to people with schizophrenia** | Good Perception | 336(71.6) | 133(28.4) | 1.00 | 1.00 |  |
|  | Poor Perception | 54(36.5) | 94(63.5) | 4.39(2.98,6.49) | 4.95(3.25,754) | **0.001\*** |
| **Wealth index** | Low | 124(61.4) | 78(38.6) | 1.11(0.75, 1.65) | 0.99(0.64,1.55) | 0.976 |
|  | Moderate | 135(64.3) | 75(35.7) | 0.98(0.66,1.47) | 0.87(0.56,1.35) | 0.524 |
|  | High | 131(63.9) | 74(36.1) | 1.00 | 1.00 |  |

**Note:** MI (mental illness), \* $p < 0.05$ ; AOR adjusted odds ratio; COR crudes odds ratio; 1.00 reference category, CI confidence interval.

## DISCUSSION

Among the total study participants 76% (95% CI: 72.6%-79.4%) have a good perception towards people with schizophrenia. However, the finding of this study is higher than the study conducted among residents of Hawassa City, South Ethiopia, 66.5%[13]. But, the finding of this study is lower than the study findings in Zaare Community[17]. This finding may imply a clue to for policymakers to establish and strengthen community-based rehabilitation for the people living with the mental health problem particularly for schizophrenic patients. Besides, it might also aid a policymaker or health professional for better effectiveness of mental health program through integrating patients care into a community where by they can live and possibly work to be able to self-independent in the community.

The magnitude of positive attitude towards people with schizophrenia among residents in Arba Minch Zuriya woread was 63.2% (95% CI: 59.4%-67.0%). This finding was higher than the study done in Southern Ethiopia, 37.3%[13]. However, the finding of this study is lower than the study findings in Greek (ref) in which the majority of the public had stigmatizing attitudes toward people living with schizophrenia. This discrepancy may be due to differences in socio-demographic and living standards and sample size difference.

Talkativeness and self-neglect behaviors were the most commonly mentioned manifestation of schizophrenia among residents in Arba Minch Zuriya woreda. This finding supported by a study conducted in Zaare Community[17]. This finding suggests that one has to exhibit behavior that draws public attention and thus socially disturbing, to be recognized as having a mental disorder. Also, health professionals should give special care to people who come with the complaint of talkativeness and self-neglect.

As the study participant mentioned that substance misuse and head injury were the most perceived cause of schizophrenia. This finding is in line with the study conducted in Zaare Community[17]. Based on this finding the policymakers should develop intervention to schizophrenia focusing on causes. The community member should keep their environment free from substance misuse and injury. This may imply that we should need to provide more knowledge and increased awareness of the causes of mental illness and also dispel the myths around the causes of the disorders.

Concerning the preferred place for the treatment of schizophrenia, the majority of the respondent mentioned spiritual/traditional places. This finding is in line with studies conducted in Agaro town, Gimbi town and Hawassa City, South Ethiopia [13,14,18]. Thus, the need for awareness sessions for the community on possible treatment options for better effectiveness of modern antipsychotic treatment is important because the majority of people in this study area seek treatment from traditional methods as a preferred method. Moreover, this study gives a hint of improvement in the accessibility of mental health services in the communities.

The current study demonstrated that youths experienced a positive attitude towards people schizophrenia more than those of older-aged years. This study is in line with the study done in Riyadh, Saudi Arabia [19]. But, this study finding not supported by the study conducted in Southern Ghana[20]. A possible reason for this may be the fact that youths are often the victim of some type of mental health disorder during their academic life. As well those youths may have more exposure to media than those of older aged.

The sex of respondents was also one of the factors associated with a positive attitude towards people with schizophrenia. Those females were more likely to experience a positive attitude towards people with schizophrenia than males. This finding was supported by studies conducted in Singapore and Southern Ethiopia [15,21]. But, this finding is inconsistent with the finding from a study conducted in Southern Ghana and Riyadh, Saudi Arabia [19,20]. The difference might be due to variation in study time and source of data.

The finding of this study found that those study participants who had no formal education and primary educated were more likely to have a positive attitude to people with schizophrenia. This finding was inconsistent with studies conducted in Riyadh, Saudi Arabia, Singapore and Southern Ethiopia [13,19,21]. The reason for this difference might be associated with living standards. Since this study was conducted in the rural part of Ethiopia, but previously conducted studies were from the urban part. Secondly, it might be due to educational access differences between urban and rural.

Furthermore, the community perception towards people with schizophrenia was one of the factors associated with a positive attitude towards people with schizophrenia. Those respondents who have poor perception towards people with schizophrenia were more likely to have a positive attitude towards people with schizophrenia.

## Limitation of the study

This study may have some limitations. First, as this study was cross-sectional study it could not ascertain the relationship between exposures and outcome. Secondly, there may be social desirability bias on information of attitude towards people living with schizophrenia since we collected the data by using interviewer administrative tools. Thirdly, we do not know whether highly symptomatic people were more or less likely to participate

## Conclusion

In conclusion, this study found that nearly half of the study participants have a negative attitude towards people with schizophrenia. Talkativeness and self-neglect behaviors were the most commonly mentioned manifestation of schizophrenia. Substance misuse and head injury were the most perceived cause of schizophrenia. Spiritual/traditional methods were preferred place for the treatment schizophrenia age of respondent, sex of respondent; educational status and perception towards people with schizophrenia were significantly associated with positive attitude towards people with schizophrenia. Therefore, giving special attention to females, youths, uneducated and individuals who have poor towards people with schizophrenia is crucial. Also, future mental health promotion activities should focus on cause and common manifestation of schizophrenia to improve the attitude toward people with schizophrenia

## Data Availability

Availability of data and materials
The data used to support the findings of this study are available from the corresponding author upon request with an email address of

## Abbreviations

AM-HDSS: Arba Minch Health and Demographic Surveillance System sites
AOR: adjusted odds ratio
CI: confidence interval
COR: crudes odds ratio
MI: mental illness
VIF: variance inflation factor

## Ethics consideration

The letter of ethical approval was obtained from the institutional review board of the College of Medicine and Health Sciences at Arba Minch University. Written consent from all participants was obtained after being fully informed about the objectives and procedures of the study for study participants. The confidentiality and privacy of participants were actively protected. All participants were assigned a unique identification number. Every effort was made to emphasize the voluntariness of this study and decisions to stop or discontinue from the study at any time.

## Acknowledgments

We would like to acknowledge all study participants for their voluntary participation in this study. We would like to extend our gratitude to Arba Minch University for all the support and opportunity provided for us to conduct this study. Last but not least, we also extend our gratitude to AM-HDSS office: for providing us the sampling frame and for their continuous support during data collection time.

## Availability of data and materials

The data used to support the findings of this study are available from the corresponding author upon request with an email address of

## Authors Contributions

Conceptualization: NB.SH, AM, and GA

Data curation: NB.SH,AM,GA,SG,TF,MK and GT

Formal analysis: NB.SH, AM, MK, and GA

Funding acquisition: NB.SH,AM,GA,SG,TF,MK and GT

Methodology: NB.SH,AM,GA,SG,TF,MK and GT

Project administration: NB, SH, and GA

Visualization: NB.SH,AM,GA,SG,TF,MK and GT

Writing – original draft: NB, SH, and GT

Writing – review & editing: NB.SH,AM,GA,SG,TF,MK and GT

